# Enhancing Infectious Intestinal Disease diagnosis through metagenomic and metatranscriptomic sequencing of over 1000 human diarrhoeal samples

**DOI:** 10.1101/2023.04.03.23288067

**Authors:** Edward Cunningham-Oakes, Blanca M. Perez-Sepulveda, Yan Li, Jay C. D. Hinton, Charlotte A. Nelson, K. Marie McIntyre, Maya Wardeh, Sam Haldenby, Richard Gregory, Miren Iturriza-Gómara, Christiane Hertz-Fowler, Sarah J. O’Brien, Nigel A. Cunliffe, Alistair C. Darby the INTEGRATE consortium

## Abstract

Fundamental issues in the traditional surveillance of diarrhoeal disease need to be addressed. The limitations of traditional microbiological diagnostic methods often mean that the cause of diarrhoea remains unknown, especially for novel or difficult-to-isolate pathogens. Sequencing samples directly, without isolating pathogens, would address this issue. However, we must ensure that sequencing also captures pathogens that are detectable using current diagnostic methods.

We show that metagenomic and metatranscriptomic approaches can effectively detect nine gastrointestinal pathogens in the UK. Metatranscriptomics shows increased sensitivity of detection for pathogens like *Campylobacter*, *Clostridioides difficile*, *Cryptosporidium* and *Giardia*, while metagenomics is more effective for detecting pathogens such as *Adenovirus*, pathogenic *Escherichia coli*, *Salmonella*, *Shigella*, and *Yersinia enterocolitica*. Certain pathogens were detected by both metagenomic and metatranscriptomic sequencing. Metatranscriptomics gave near-complete genome coverage for Human mastadenovirus F and detected *Cryptosporidium* via capture of *Cryptosporidium parvum* virus (CSpV1). A comprehensive transcriptomic profile of *Salmonella* Enteritidis was recovered from the stool of a patient with a laboratory-confirmed *Salmonella* infection.

This study highlights the power of direct sequencing of human samples to augment GI pathogen surveillance and clinical diagnostics. Metatranscriptomics was best for capturing a wide breadth of pathogens and was more sensitive for this purpose. We propose that metatranscriptomics should be considered for future surveillance of gastrointestinal pathogens. This study has generated a rich data resource of paired metagenomic and metatranscriptomic datasets, direct from over 1000 patient stool samples. We have made these data publicly available to promote the improved understanding of pathogens associated with infectious intestinal diseases.

## Background

The incidence of infectious intestinal disease (or acute gastroenteritis) is estimated to be 18 million cases each year in the United Kingdom (UK)[1]. About 25% of infected people experience diarrhoeal and related gastrointestinal symptoms. The current mainstay for identifying gastrointestinal pathogens in faecal specimens in the UK are conventional laboratory techniques, including microscopy and antigen detection, and increasingly, molecular assays such as nucleic acid amplification[2].

Although conventional and polymerase chain-reaction (PCR)-based approaches (such as BioFire Panels) are validated for clinical laboratory use[2], both focus on a single gene or set of characteristics providing limited information about pathogens[3]. In the case of bacterial culture, the time required for growth, lack of sensitivity, and the challenge of culturing fastidious organisms cause diagnostic delays[3]. Current methods lack the sensitivity required to detect pathogens that are present intermittently or in low numbers[4]. In contrast, PCR-based methods use target sequences for organism detection, resulting in increased sensitivity and no strict requirement for the prior growth of organisms[3].

Whilst PCR-based methods are more sensitive than conventional (traditional) methods, they lack resolution[5] and are unable to achieve the strain-level discrimination required for outbreak monitoring[6]. Inevitably, molecular assays target known genes from well-characterised organisms [5], meaning that unexpected pathogens and unique genes will be missed. Whole-genome sequencing partly overcomes this, but still requires the isolation of a pure culture of pathogenic organisms.

The speed and sensitivity of metagenomic and metatranscriptomic data analysis[7] has been significantly enhanced by k-mer-based methods, an approach that has been widely adopted in many popular workflows[8,9] to identify pathogens in metagenomic samples through database matching. The computational efficiency of k-mers is ideal for high-throughput sequencing applications[10]. However, it is important to note that sequencing errors and the comprehensiveness of the databases used[11] can influence the effectiveness of k-mer-based approaches.

It has been proposed that DNA and RNA sequencing of clinical samples could be a valuable future approach[5]. To ensure that the presence of pathogens is recorded accurately, it is essential to understand the strengths and limitations of metagenomic and metatranscriptomic approaches.

The INTEGRATE study[12] compared traditional (culture, ELISA, microscopy and PCR) diagnostic methods with state-of-the-art, sensitive molecular and genome-based microbiological methods for identifying and characterising causative pathogens[12]. Here, we present data generated by next-generation sequencing of the stool microbiomes of 1,067 patients with symptoms of gastroenteritis. This dataset represented a unique opportunity to explore the effectiveness of k-mer-based analyses using a large number of samples that were also characterised with gold-standard clinical laboratory diagnostics. We considered the comparative benefits of different sequencing types in various scenarios (right test, right time, right patient).

We use these data to show that both metagenomic (DNA) and metatranscriptomic (RNA) sequencing directly from stool can detect the major community-associated gastrointestinal (GI) pathogens in the United Kingdom. We found that metagenomic and metatranscriptomic sequencing have distinctive features for pathogen detection, and discovered that metatranscriptomics offers unexpected benefits for pathogen surveillance.

All of these data have been made publicly available (PRJEB62473) to provide a rich data source for researchers to foster a deeper understanding of the pathogens associated with infectious intestinal diseases.

## Results

### Metagenomics and metatranscriptomics show different levels of sensitivity for GI pathogens

The DNA and RNA from a total of 1,067 samples were sequenced, with 985 providing both metagenomic and metatranscriptomic data (see Supplementary File 1 for all k-mer counts and associated taxonomy from these samples). For *Campylobacter*, *Cryptosporidium*, and *Giardia* (Figure 1), metatranscriptomics showed greater sensitivity than metagenomics (see Methods for definition of sensitivity). In contrast, metagenomics displayed greater sensitivity than metatranscriptomics for the *Adenoviridae*, *Clostridium difficile*, pathogenic *Escherichia coli, Salmonella, Shigella* and *Yersinia enterocolitica* (Figure 1). *Entamoeba histolytica* were not detected in either the metagenomic or metatranscriptomic datasets.

**Figure 1:**
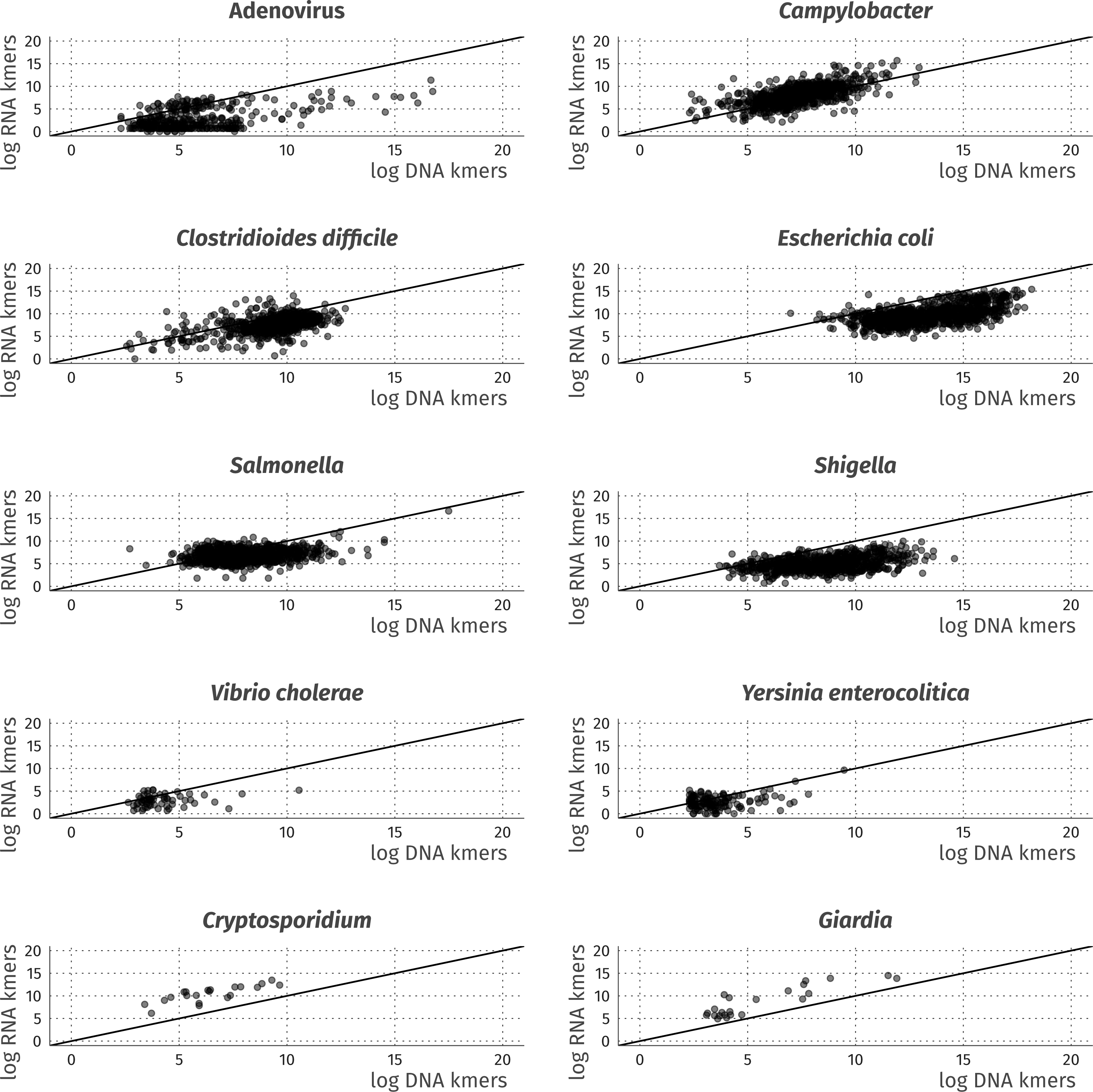
Visual overview and comparison of DNA (metagenomic) and RNA (metatranscriptomic) sequencing reads. assigned to GI pathogens of relevance to the UK setting. For all graphs, the dashed (black) intercept line is provided to highlight the skew of sensitivity towards either DNA or RNA.

### The detection of GI pathogens in metagenomic and metatranscriptomic data mirrors clinical laboratory results

Our analysis showed that the pathogens detected in sequencing reads closely match results generated by laboratory diagnostics for Adenovirus, *C. difficile*, *Campylobacter*, *Cryptosporidium*, Norovirus, Rotavirus, *Salmonella*, Sapovirus*, Shigella,* and *Y. enterocolitica* (Figure 2). Most major GI community pathogens in the United Kingdom were detected in both metagenomic and metatranscriptomic data, but RNA viruses could only be detected by metatranscriptomics. A summary of the “traditional” methods used for pathogen diagnosis in the INTEGRATE study is presented in Table 1.

**Figure 2:**
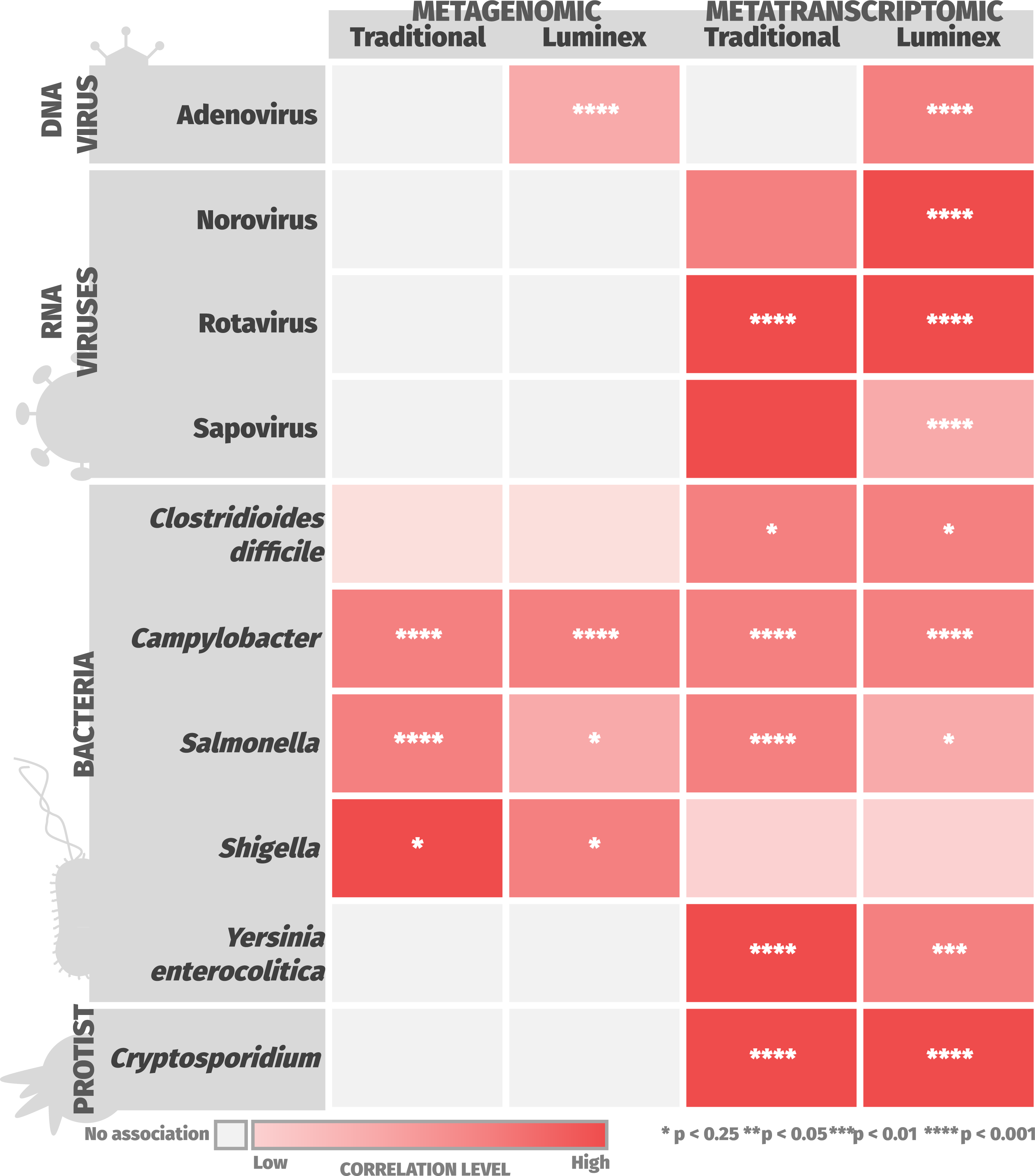
Statistically significant correlations were observed between sequencing data and laboratory tests for 10 out of 14 major GI community pathogens in the United Kingdom. Results where at least one statistically significant correlation was observed are shown. All correlations, whether significant or not, are displayed in Supplementary Figure 1. No statistically significant correlation was found between the sequencing and diagnostic test for Astrovirus, *E. histolytica*, *Giardia* or *V. cholerae*. The darker the colour of a quadrant in a heatmap, the stronger the correlation (coefficient) between the detection of a pathogen in sequencing data (metagenomic or metatranscriptomic) and a laboratory result (Luminex or Traditional). Asterisks in quadrants indicate the statistical significance of correlations as follows: *: p < 0.25; **: p < 0.05; ***: p < 0.01; ****: p < 0.001.

**Table 1:**
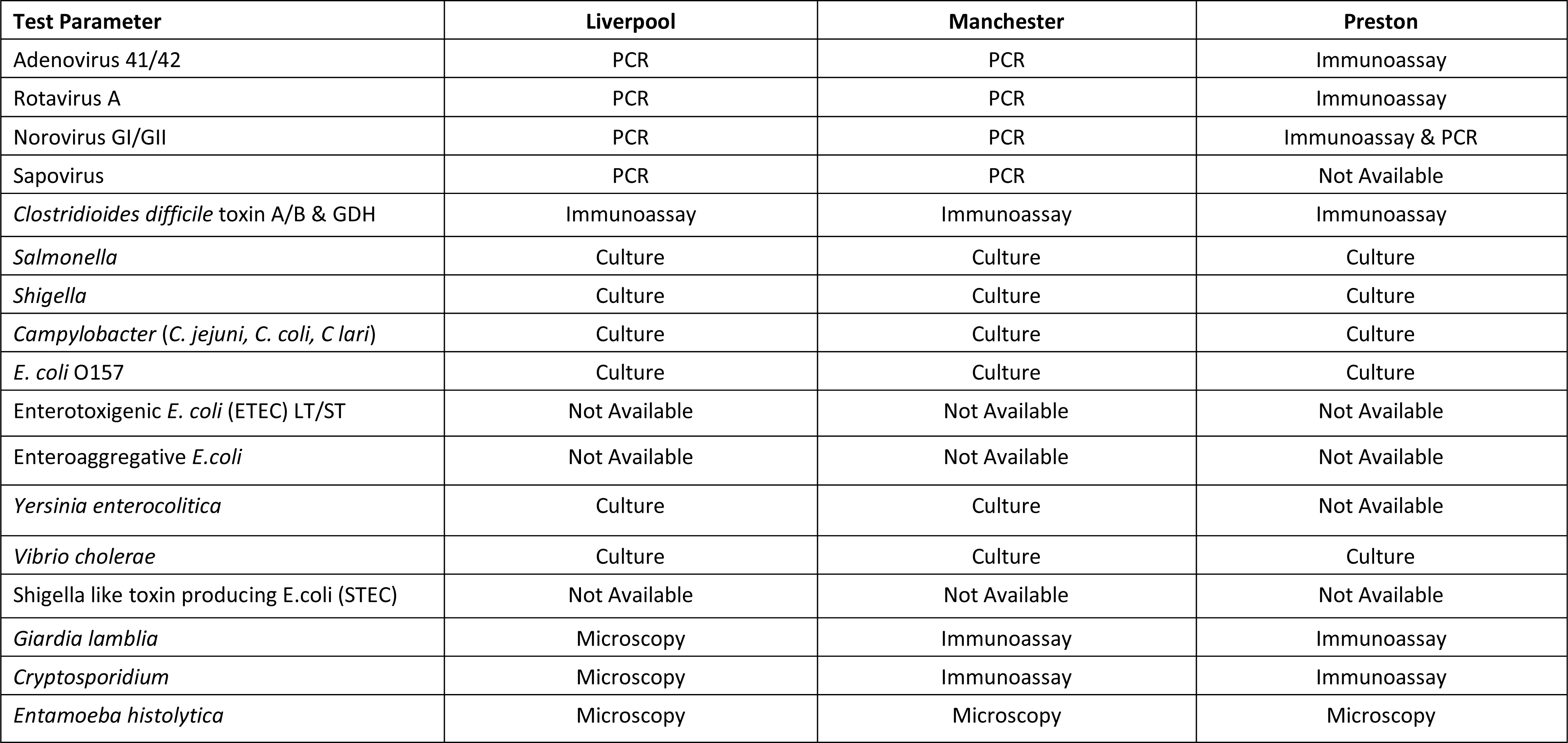
Summary of “traditional” methods used at clinical laboratories during the INTEGRATE study. Samples were processed via routine diagnostic pathways at each laboratory involved in the study (see Supplementary File 5). Traditional assays for Enterotoxic and Enteroaggregative *E. coli*, as well as *E. coli* O157, were not available (only available in the Luminex xTAG GPP panel).

| Test Parameter | Liverpool | Manchester | Preston |
| --- | --- | --- | --- |
| Adenovirus 41/42 | PCR | PCR | Immunoassay |
| Rotavirus A | PCR | PCR | Immunoassay |
| Norovirus GI/GII | PCR | PCR | Immunoassay & PCR |
| Sapovirus | PCR | PCR | Not Available |
| *Clostridioides difficile* toxin A/B & GDH | Immunoassay | Immunoassay | Immunoassay |
| *Salmonella* | Culture | Culture | Culture |
| *Shigella* | Culture | Culture | Culture |
| *Campylobacter* (*C. jejuni*, *C. coli*, *C. lari*) | Culture | Culture | Culture |
| *E. coli* O157 | Culture | Culture | Culture |
| Enterotoxigenic *E. coli* (ETEC) LT/ST | Not Available | Not Available | Not Available |
| Enteraggregative *E. coli* | Not Available | Not Available | Not Available |
| *Yersinia enterocolitica* | Culture | Culture | Not Available |
| *Vibrio cholerae* | Culture | Culture | Culture |
| Shigella like toxin producing *E. coli* (STEC) | Not Available | Not Available | Not Available |
| *Giardia lamblia* | Microscopy | Immunoassay | Immunoassay |
| *Cryptosporidium* | Microscopy | Immunoassay | Immunoassay |
| *Entamoeba histolytica* | Microscopy | Microscopy | Microscopy |

### Viral pathogens

DNA viruses such as Adenovirus were detected in both the metagenomic and metatranscriptomic datasets. For Adenovirus, positive correlations were observed between detection in metagenomic reads, metatranscriptomic reads, and Luminex xTAG Gastrointestinal Pathogen Panel (Luminex) results (p<0.001). The metatranscriptomic results correlated positively with Rotavirus (p<0.001). The detection of Norovirus and Sapovirus by metatranscriptomics was significantly correlated (p<0.001) with the Luminex results. Metagenomic and metatranscriptomic results did not correlate with the detection of Astrovirus using Traditional and Luminex methods.

### Protists

Protists were detected by both metagenomics and metatranscriptomics. However, the metatranscriptomic results had a much higher sensitivity for the detection of parasites than metagenomics. Positive correlations between the detection of *Cryptosporidium* in metatranscriptomic data and laboratory data were highly significant (p<0.001). No associations were observed between metagenomic data and laboratory results for *Cryptosporidium*. There was no correlation between the detection of *Giardia* using Traditional or Luminex methods, and detecting *Giardia* using metagenomics or metatranscriptomics.

### Bacterial pathogens

The identification of bacterial pathogens from sequencing data is challenging, as commensal organisms and pathogens can have extremely high levels of genomic similarity. Laboratory diagnostics tend to differentiate commensal and pathogenic organisms using genes or phenotypes associated with pathogenicity. Our results show that metagenomics and metatranscriptomics can both identify bacterial pathogens with differing sensitivities. For *Campylobacter*, positive correlations were observed between direct sequencing and all laboratory results (p<0.001). *Salmonella* displayed positive correlations between sequencing data and both Traditional (p<0.001) and Luminex (p<0.25) diagnostics. *C. difficile* metatranscriptomic sequencing data positively correlated with both Traditional (p<0.25) and Luminex (p<0.001) diagnostics. *Y. enterocolitica* sequencing data positively correlated with Luminex results as follows: *C. difficile* metatranscriptomic reads (p<0.001), *Y. enterocolitica* metatranscriptomic reads (p<0.01), and *Y. enterocolitica* metagenomic reads (p<0.001).

*E. coli* and *Shigella* are closely-related species; detection of *Shigella* in metagenomic data correlated positively with traditional and Luminex diagnostics (p<0.25), while *E. coli* showed a non-significant correlation (p>0.25). *Vibrio cholerae* were not be detected in either metagenomic or metatranscriptomic data, consistent with laboratory diagnostics, which identified no *V. cholerae* infections.

A summary of all correlations between the detection of GI pathogens in sequencing reads and laboratory data and their significance is provided in Supplementary Files 2 and 3.

### Case-studies for the use of metatranscriptomics in pathogen surveillance

#### Complete genomes from diarrhoeal-associated Adenovirus can be detected in both metagenomic and metatranscriptomic data

Whilst Adenovirus is a DNA virus, it was surprising to see that Adenovirus could also be detected in RNA. There was a strong correlation between the detection of Adenovirus in metagenomic and metatranscriptomic data, and detection using Luminex methods (see Supplementary File 2 and Figure 2). Mapping of Adenovirus-associated reads to the Human mastadenovirus F genome showed that, in 7 out of 9 samples, metagenomics generated more complete genes, at a higher depth (Supplementary File 4). However, in 2 out of 9 samples (Samples 5638 and 6985) near-complete genome coverage was achieved using both metagenomics and metatranscriptomics (83.9-100% - Supplementary File 4 and Figure 3). These results demonstrate the potential of metatranscriptomics to directly capture the virome from clinical samples, including DNA viruses relevant to the condition of interest.

**Figure 3:**
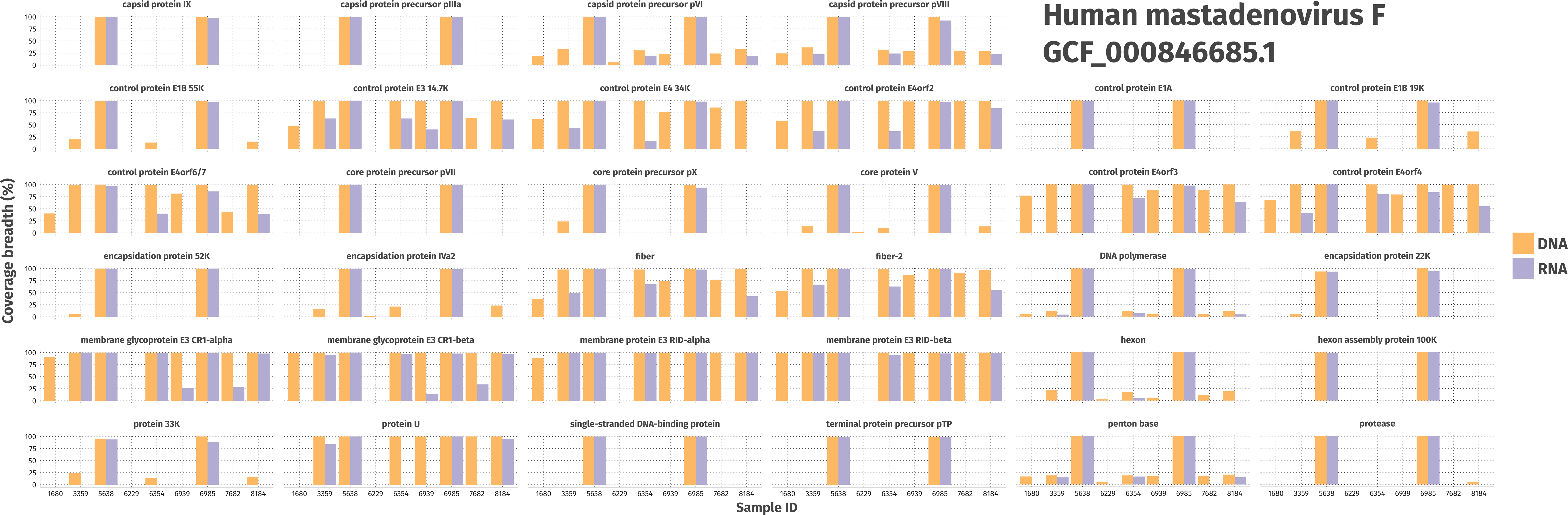
Adenovirus can be detected through its genomic material and the expression of transcript, directly from stool. Graphs display the breadth of coverage (%) for both DNA (purple) and RNA (orange) across nine samples, chosen on the basis of positive results through gold-standard laboratory methods. Coverage values were generated via mapping to a representative Human mastadenovirus F genome (GCF_000846685.1, International Committee on Taxonomy of Viruses species exemplar).

#### Cryptosporidium-associated RNA viruses facilitate detection directly from stool

Another interesting observation was the correlation (p<0.001, see Figure 2) between the detection of *Cryptosporidium* using metatranscriptomics and the detection of *Cryptosporidium* in the laboratory. In contrast, detecting *Cryptosporidium* using metagenomics did not correlate with laboratory results. Mapping revealed that *Cryptosporidium* was accurately identified in metatranscriptomic data due to the presence of *Cryptosporidium parvum* virus (CSpV1), which is an RNA virus. CSpV1 was identified in 33 metatranscriptomic samples (Table 2). Of these 33 samples, 9 received a positive result using Traditional methods, whilst 16 were positive by Luminex. CSpV1 received a high-confidence score (0.995) in 21 out of the 33 samples, with percentage breadth of genome coverage ranging from 57.1-100%. This illustrates the potential of CSpV1 to be used as a reliable biomarker for human *Cryptosporidium* infection.

**Table 2:**
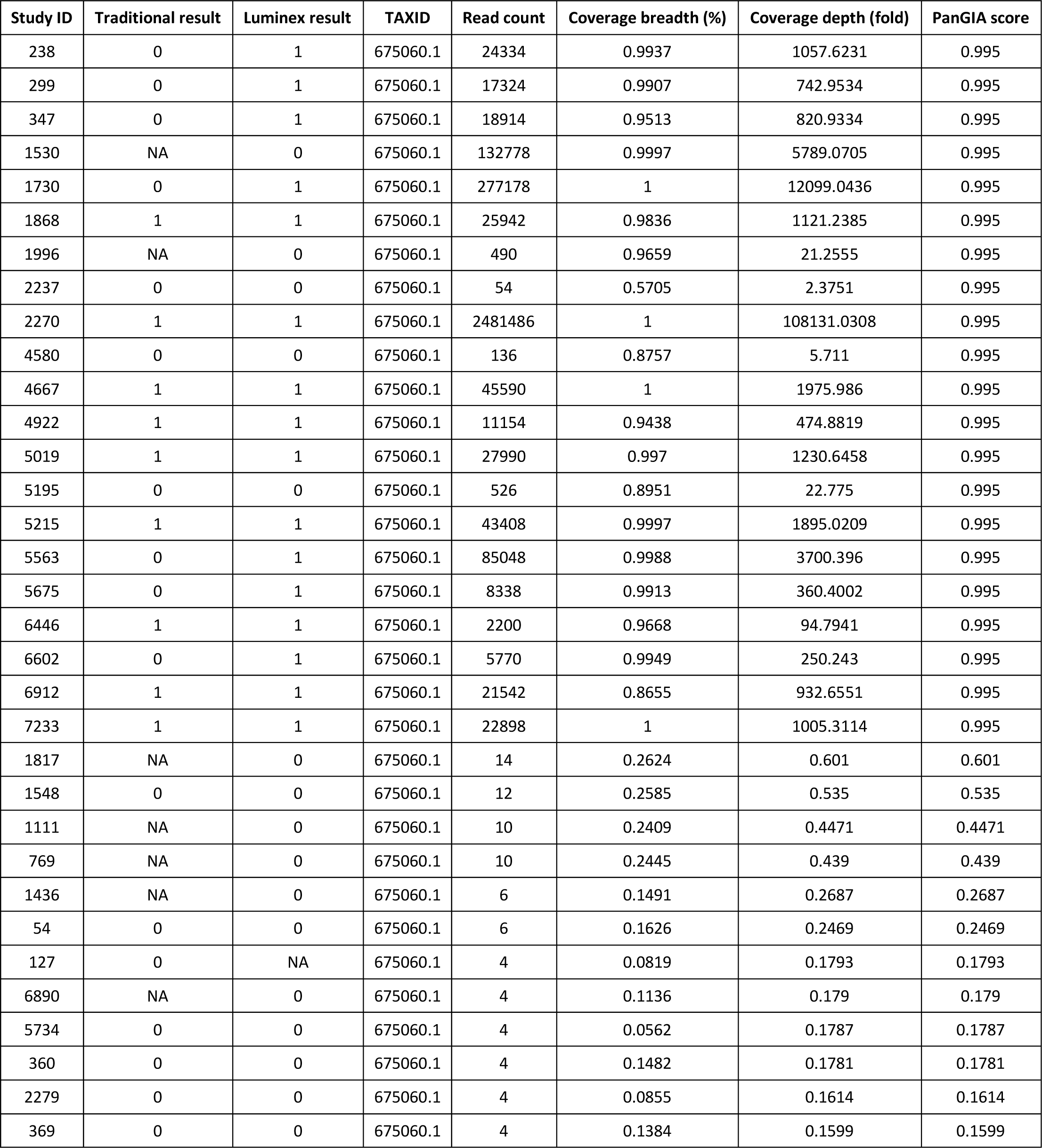
Identification of CSpV1 in metatranscriptomic data in comparison to results from *Cryptosporidium* laboratory diagnostics. For both Traditional and Luminex results, NA represents instances where a diagnostic test could not be performed.

| Study ID | Traditional result | Luminex result | TAXID | Read count | Coverage breadth (%) | Coverage depth (fold) | PanGIA score |
| --- | --- | --- | --- | --- | --- | --- | --- |
| 238 | 0 | 1 | 675060.1 | 24334 | 0.9937 | 1057.6231 | 0.995 |
| 299 | 0 | 1 | 675060.1 | 17324 | 0.9907 | 742.9534 | 0.995 |
| 347 | 0 | 1 | 675060.1 | 18914 | 0.9513 | 820.9334 | 0.995 |
| 1530 | NA | 0 | 675060.1 | 132778 | 0.9997 | 5789.0705 | 0.995 |
| 1730 | 0 | 1 | 675060.1 | 277178 | 1 | 12099.0436 | 0.995 |
| 1868 | 1 | 1 | 675060.1 | 25942 | 0.9836 | 1121.2385 | 0.995 |
| 1996 | NA | 0 | 675060.1 | 490 | 0.9659 | 21.2555 | 0.995 |
| 2237 | 0 | 0 | 675060.1 | 54 | 0.5705 | 2.3751 | 0.995 |
| 2270 | 1 | 1 | 675060.1 | 2481486 | 1 | 108131.0308 | 0.995 |
| 4580 | 0 | 0 | 675060.1 | 136 | 0.8757 | 5.711 | 0.995 |
| 4667 | 1 | 1 | 675060.1 | 45590 | 1 | 1975.986 | 0.995 |
| 4922 | 1 | 1 | 675060.1 | 11154 | 0.9438 | 474.8819 | 0.995 |
| 5019 | 1 | 1 | 675060.1 | 27990 | 0.997 | 1230.6458 | 0.995 |
| 5195 | 0 | 0 | 675060.1 | 526 | 0.8951 | 22.775 | 0.995 |
| 5215 | 1 | 1 | 675060.1 | 43408 | 0.9997 | 1895.0209 | 0.995 |
| 5563 | 0 | 1 | 675060.1 | 85048 | 0.9988 | 3700.396 | 0.995 |
| 5675 | 0 | 1 | 675060.1 | 8338 | 0.9913 | 360.4002 | 0.995 |
| 6446 | 1 | 1 | 675060.1 | 2200 | 0.9668 | 94.7941 | 0.995 |
| 6602 | 0 | 1 | 675060.1 | 5770 | 0.9949 | 250.243 | 0.995 |
| 6912 | 1 | 1 | 675060.1 | 21542 | 0.8655 | 932.6551 | 0.995 |
| 7233 | 1 | 1 | 675060.1 | 22898 | 1 | 1005.3114 | 0.995 |
| 1817 | NA | 0 | 675060.1 | 14 | 0.2624 | 0.601 | 0.601 |
| 1548 | 0 | 0 | 675060.1 | 12 | 0.2585 | 0.535 | 0.535 |
| 1111 | NA | 0 | 675060.1 | 10 | 0.2409 | 0.4471 | 0.4471 |
| 769 | NA | 0 | 675060.1 | 10 | 0.2445 | 0.439 | 0.439 |
| 1436 | NA | 0 | 675060.1 | 6 | 0.1491 | 0.2687 | 0.2687 |
| 54 | 0 | 0 | 675060.1 | 6 | 0.1626 | 0.2469 | 0.2469 |
| 127 | 0 | NA | 675060.1 | 4 | 0.0819 | 0.1793 | 0.1793 |
| 6890 | NA | 0 | 675060.1 | 4 | 0.1136 | 0.179 | 0.179 |
| 5734 | 0 | 0 | 675060.1 | 4 | 0.0562 | 0.1787 | 0.1787 |
| 360 | 0 | 0 | 675060.1 | 4 | 0.1482 | 0.1781 | 0.1781 |
| 2279 | 0 | 0 | 675060.1 | 4 | 0.0855 | 0.1614 | 0.1614 |
| 369 | 0 | 0 | 675060.1 | 4 | 0.1384 | 0.1599 | 0.1599 |

#### Generation of a complete transcriptomic profile for Salmonella

Metatranscriptomic analysis of stool from a patient with a laboratory-confirmed *Salmonella* infection yielded functional insights that cannot be achieved with Traditional and Luminex diagnostics. Transcriptomic analysis reveals gene expression patterns that key biological processes in bacteria. The high-quality transcriptomic profile was generated from 12.7 million sequence reads that mapped to the genome of *S. enterica* serovar Enteritidis PT4 strain P125109. The *S.* Enteritidis transcripts from this novel gene expression data can be visualised and interrogated in a bespoke genome browser (https://s.hintonlab.com/study_74).

A variety of environmentally responsive *Salmonella* genes were highly expressed (as defined by Kröger *et al*. 2013; *Cell Host Microbe* [13]), likely reflecting physicochemical stresses the bacteria had been exposed to in the stool sample. Examples include *ahpC* (oxidative stress), *hmpA* (nitrosative stress), *phoH* (phosphate starvation), *pspA* (extracytoplasmic stress), and the *rpoE* and *rpoS* transcription factor genes, as can be seen with the [SalComMac data visualisation tool](https://bioinf.gen.tcd.ie/cgi-bin/salcom.pl?header_rotation=45;query=ahpC%20hmpA%20phoH%20pspA%20rpoE%20rpoS;db=salcom_mac_HL). The unexpected discovery that the metatranscriptomic analysis of a human stool sample can generate a comprehensive gene expression profile of a *Salmonella* pathogen is worthy of future exploitation.

## Discussion

We have demonstrated that metagenomic and metatranscriptomic approaches provide agnostic detection of important UK GI pathogens from human stool. The primary impact of this work lies within GI pathogen diagnostics. Our findings demonstrate the potential for improving current GI pathogen diagnostics and the bridging of gaps not addressed by standard approaches.

### Improvements within the scope of current diagnostics

Sequencing directly from stool could minimise the time required for pathogen detection, allowing more laborious detection methods such as cultivation to be appropriately tailored to confirm the presence of the suspected pathogens.

The metatranscriptomic strategy displays increased sensitivity for *Campylobacter*, *C. difficile, Cryptosporidium* and *Giardia*, whilst metagenomics displayed increased sensitivity for other GI pathogens including Adenovirus, pathogenic *E. coli*, *Salmonella*, *Shigella,* and *Y. enterocolitica*. Direct extraction of RNA from stool represents a single sample format and cultivation-independent process for detecting a broad range of GI pathogens, including unexpected aetiological agents and those that cannot be detected by metagenomic sequencing, such as RNA viruses. The observation of near-complete genome coverage for Human mastadenovirus F in both the metagenome and metatranscriptome highlights the potential to optimise metatranscriptomic sequencing from stool to capture the virome, including DNA virus transcriptomes relevant to clinical conditions. This finding is supported by previous clinical studies, which used metatranscriptomics to simultaneously measure the virome, microbiome, and host response[14]. Our data and previous studies[15] confirm the ability to characterise disease-related microbiomes with increased sensitivity via metatranscriptomics.

Increased sensitivity for the detection of protists of concern in GI infections was also demonstrated. Our visualisations of metagenomic and metatranscriptomic reads (Figure 1) showed that metatranscriptomic data provide greater sensitivity for detecting *Cryptosporidium* and *Giardia* (protists). Finally, our multivariable model demonstrated the strong correlation and high significance between the detection of *Cryptosporidium* in the laboratory, and in metatranscriptomic data, a finding that was supported by a previous study that detected 23% more blood infections than traditional methods[16]. In these data, the presence of *Cryptosporidium*-associated viruses increases the sensitivity of detection for this particular protist. In contrast, the detection of *Cryptosporidium* using metagenomics appears to be spurious. This virus has recently been reported in various subtypes of *Cryptosporidium parvum* from diarrhoeic farm animals[17,18], but it is not currently used as a diagnostic marker in humans. These results highlight the advantages of metatranscriptomics for *Cryptosporidium* surveillance, where the use of metagenomics alone could result in missed identification. This suggests that RNA viruses could be considered sensitive biomarkers for *Cryptosporidium* and other protists.

Overall, our findings reveal that RNA is a valuable diagnostic target for the detection of pathogens of low abundance and reduces false-positive signals from commensals. Our approach could influence the future allocation of resources for reference laboratory diagnostics.

### Bridging gaps not addressed by current diagnostics

Metatranscriptomic data could fill gaps in areas of clinical relevance that are not fulfilled by routine clinical diagnostics. Firstly, metagenomic and metatranscriptomic data permits the identification of multiple species and strains within a sample (Supplementary Figure 1, Supplementary File 4), including novel pathogens. Such analysis is beyond the scope of our study, but has been used to successfully identify novel pathogens from the stools of various mammalian species[19,20]. Additionally, we have demonstrated the ability to rapidly generate gene expression profiles for pathogens of concern, without prior enrichment. Finally, we have generated illuminating metatranscriptomic data from a human diarrhoeal sample. Future studies could generate true disease-state expression profile by using appropriate methodology. From a clinical perspective, the use of metagenomic and metatranscriptomic sequencing has the potential to reveal the effects of interventions[21] and to accurately investigate host-pathogen dynamics during genuine human infections[14].

### Limitations

In certain scenarios, metagenomic sequencing captures more information than metatranscriptomic sequencing. For DNA viruses, while it is possible to capture expression profiles, optimisation is needed to improve this process. Our data demonstrate that the underlying biology can be captured, but further refinement is necessary. Additionally, *E. histolytica* was not captured by metagenomic or metatranscriptomic approaches, a finding that requires further investigation.

Future adaptation of our workflow is needed for the accurate identification of *E. coli* pathovariants from sequencing data. *Shigella* and *E. coli* pathovariants are extremely similar on a genome-wide (and taxonomic) level[22], and are currently distinguished using specific gene-based assays[23]. In contrast, our study drew correlations between pathogens in reads and laboratory tests based on taxonomy. Due to this approach, and the ubiquitous presence of *E. coli* in all stool samples, it was not possible to associate the presence of *E. coli*, and gene-based assays used for *E. coli* pathovariant identification (Supplementary Table S1, Supplementary File 4). This may explain the limited overlap between *Shigella* sequencing reads and laboratory tests (Figure 2), due to the conflation of *Shigella* with *E. coli* (and *vice versa*) in our current analysis. Future work should also validate this approach on a range of sample types (beyond stool) to ensure robustness and reliability across different clinical scenarios.

### Perspective

With sufficient benchmarking, the diagnosis of various GI pathogens can be achieved from clinical samples without culturing. Metatranscriptomics can detect active DNA viruses and enhance sensitivity for protists by using RNA viruses as biomarkers. Perhaps the value of clinical metagenomics has been overstated, and metatranscriptomics could offer a comprehensive approach to both detect disease-relevant pathogens and understand their biology.

To our knowledge, this study is the first to demonstrate and quantify the benefits that metatranscriptomics could bring to gastrointestinal surveillance in the United Kingdom by direct comparison of all major community pathogens to validated diagnostics. Our study lays the groundwork for the implementation of sequencing-based diagnostics in clinical settings, with the potential to detect a broader range of organisms than current approaches and to identify novel pathogens.

## Materials and methods

### Patient recruitment and sample collection

Recruitment and sample collection was described previously[12]. Briefly, stool was collected from 1,067 members of the public with symptoms of acute gastroenteritis via practices in the Royal College of General Practitioners Research and Surveillance Centre National Monitoring Network (RCGP RSC NMN). Patients meeting inclusion criteria were invited to submit a stool sample for microbiological analysis. Consent was obtained for this procedure, as stool sampling is usually only performed if a case is severe, or persistent. Patients who provided a stool sample were then recruited into the study.

### Sample processing

Faecal samples were received by one of three clinical laboratories (Royal Liverpool and Broadgreen University Hospitals NHS Trust, Central Manchester University Hospitals NHS Foundation Trust, or Lancashire Teaching Hospitals NHS Foundation Trust), and divided into two aliquots. One part of the sample was processed using traditional methods (culture, ELISA, microscopy or PCR with no additional hybridisation probe – see Supplementary File 5) at each laboratory; the other was processed using a combined molecular multiplex real-time polymerase chain reaction (PCR) and target-specific hybridisation probe [Luminex xTAG Gastrointestinal Pathogen Panel, Luminex, I032C0324], supplemented with targets for Enteroaggregative *Escherichia coli* and Sapovirus. Nucleic acid extraction from faeces was performed using QIASymphany and EasyMag automated nucleic acid extraction platforms. Further details can be found in the primary study protocol[12]. Samples that returned a positive result according to routine clinical practice were designated as “clinical positive”. Those that returned a positive result by Luminex were designated as “molecular positive”.

### Metagenomic and metatranscriptomic sequencing

Illumina fragment libraries from DNA were prepared using NEBNext DNA Ultra kits. For the generation of dual-indexed, strand-specific RNASeq libraries, RiboZero rRNA depleted RNA samples were prepared using NEBNext Ultra Directional RNA kits. For all libraries, paired-end, 150-bp sequencing was then performed on an Illumina HiSeq 4000, generating data from >280 million clusters per lane.

### Quality control for second-generation sequencing reads

Modules from the MetaWRAP[24] (v1.3.2) pipeline were used to standardise metagenome analysis. The pipeline was deployed in a dedicated Conda environment, using the “manual installation” guide (see [GitHub](https://github.com/bxlab/metaWRAP)). All paired-end reads underwent quality-control using the MetaWRAP “read_qc” module to remove low-quality, adapter, and human sequence reads. The T2T consortium complete human genome, (GCF_009914755.1) and human mitochondrial genome (NC_012920.1) were used as references for the removal of human reads.

### Assigning taxonomy to genomic DNA and RNA reads and assessing microbiome diversity

DNA and RNA reads were used for taxonomic assignments with Kraken2[8] (v2.1.2), using a custom database, which included all RefSeq complete genomes and proteins for archaea, bacteria, fungi, viruses, plants, protozoa, as well as all complete RefSeq plasmid nucleotide and protein sequences, and a false-positive minimised version of the NCBI UniVec database.

A confidence threshold of 0.1 was set for read assignments, and reports were generated for downstream biom file generation. For DNA sequencing data, read counts assigned to taxonomies in each sample were then re-estimated using the average read length of that sample, using Bracken[25] (v2.0). Kraken-biom (v1.0.1) was then used to generate biom file in json format, using initial Kraken reports for RNA samples, and Bracken reports for DNA samples. Biom (v2.1.6) was then used to assign tabulated metadata to this biom file.

### Visualisation and comparison of taxa of interest in RNA and DNA

A taxonomy table was generated from the biom file in R (v4.2.2) using Phyloseq[26] (v1.42.0) and MicrobiotaProcess[27] (v1.10.3). Read-assigned taxonomy counts were parsed from this table for any samples with both metagenomic (DNA) and metatranscriptomic data (n=985). Counts were extracted for the following taxa: *Adenoviridae*, *Campylobacter*, *Clostridioides difficile* (*C. difficile*), *Cryptosporidium*, *Escherichia coli* (*E. coli*), Norovirus, Rotavirus, *Salmonella*, *Shigella*, Sapovirus, *Vibrio cholerae (V. cholerae*) and *Yersinia enterocolitica* (*Y. enterocolitica*). These taxa were chosen to reflect the pathogen panels used during this study. RNA virus (Astrovirus, Norovirus, Rotavirus, and Sapovirus) read counts could not be extracted for this part of the analysis, as visualisations relied on the presence of DNA reads. DNA and RNA counts were log-transformed and plotted against one another as a line graph using standard functions in ggplot2[28] (v3.4.0). Visualisations were then used to assess the sensitivity of metagenomics and metatranscriptomics for the selected taxa, where we define sensitivity as the skew of data points towards either metagenomics (x-axis) or metatranscriptomics (y-axis). A 0,0 intercept line was included in each line graph to assist in illustrating sensitivity differences.

### Correlation of genomic reads assigned to taxa of interest with number of observed taxa and results from laboratory diagnostics

Associations between read counts and laboratory results for organisms of interest were assessed using a multivariable linear regression model in MaAsLin 2[29] (v1.6.0) under default settings. The introduction of another variable into the model (laboratory results) provided a point of reference. This allowed us to determine the relationship between any sample with sequencing data and laboratory results. As such, for this analysis, all sequenced patient samples (n = 1,067) were used, even if they did not contain both metagenomic and metatranscriptomic data. Our approach allowed RNA virus read counts from metatranscriptomic data to be included in this analysis. To visualise the strength of correlations between laboratory results and pathogen-assigned sequencing reads, correlation coefficients and adjusted p-values from the model were tabulated and used to generate a heatmap with corrplot (v0.9.2). Adjusted p-values were generated using the Benjamini-Hochberg Procedure.

### Comparative analysis of Adenovirus-associated k-mers in DNA and RNA

The extract_kraken_reads.py utility from Kraken-tools (v1.2) was used alongside Kraken2 reports to extract reads with k-mer profiles associated with the family *Adenoviridae* for samples that tested positive using either traditional or Luminex methods. Samples where sequencing was not successful for both DNA and RNA were excluded from this analysis.

These reads were then mapped to the Human Mastadenovirus F genome (Accession: GCF_000846685.1) using HISAT2[30] (v2.2.1) for splice-aware mapping. Coverage statistics were then generated using samtools coverage. Coverage statistics for each sample were compiled into a single table and visualised as a bar chart using ggplot2 (v3.4.0).

### Identification of CSpV1 as a biomarker of Cryptosporidium infection

To understand why *Cryptosporidium*-associated k-mers showed a positive correlation with using gold-standard diagnostics in metatranscriptomic but not metagenomic sequencing data, we employed competitive mapping using PanGIA[31] (v1.0.0-RC6.2). We mapped quality-controlled reads from all INTEGRATE samples against a database containing representative and reference genomes of bacteria, archaea, and viruses in NCBI RefSeq (release 89). This helped to validate our k-mer-based results and offers a less computationally intensive alternative to mapping-based approaches for future users of k-mer-based databases. By aligning the reads to these genome sequences, we obtained a read count and depth of coverage for each organism. We then extracted entries associated with the term ‘*Cryptosporidium*’ along with their corresponding scores and mapping information. PanGIA also accounts for many reads mapped equally well to other organisms and the percentage of identity of these hits and derived a confidence score from this, ranging from 0 to 1 for each query sequence at each taxonomy level. This allowed us to determine the certainty that the organism is truly present in the sequencing data.

### Visualisation of a Salmonella transcriptome directly from stool

Metatranscriptomic reads from a sample of a patient with a later-confirmed (culture positive) *Salmonella* spp. infection underwent quality control, alignment, and quantification using the Bacpipe RNA-seq processing pipeline (v0.6.0). The GFF annotation[32] for the *Salmonella enterica* subsp. *enterica* serovar Enteritidis PT4 strain P125109 (Accession: GCA_015240635.1) was used in this analysis. Coverage tracks and annotation were visualised using JBrowse (v1.16.8). This visualisation can be found here: https://s.hintonlab.com/study_74.

## Declarations

### Ethics approval and consent to participate

Members of the public with symptoms of acute gastroenteritis, including a case definition of vomiting and diarrhoea, who sought health advice from general practices in the RCGP RSC NMN were invited to submit a stool sample for microbiological examination. Their consent for this procedure was sought because normal care would not necessarily entail stool sampling for most patients unless their symptoms were severe or had persisted for a long time. The North West - Greater Manchester East Research Ethics Committee (REC reference: 15/NW/0233) and NHS Health Research Authority (HRA) Confidential Advisory Group (CAG) (CAG reference: 15/CAG/0131) granted a favourable ethics opinion for the INTEGRATE project. Approval was also granted by NHS Research Management and Governance Committees (including Royal Liverpool and Broadgreen University Hospital Trust, Lancashire Teaching Hospitals NHS Foundation Trust, Central Manchester University Hospitals NHS Foundation Trust, and the University of Liverpool Sponsor), the Lancaster University Faculty of Health and Medicine Ethics Committee, and the University of Liverpool Ethics Sub-Committees. An Information Governance Toolkit (IGT) from the Department of Health hosted by the Health and Social Care Information Centre (HSCIC) was also completed for the project, and all project research staff obtained Honorary NHS contracts, research passports, and letters of access, as necessary.

### Consent for publication

The publication was approved by the National Institute for Health and Care Research on 29^th^ March 2023.

### Availability of data and materials

Illumina sequence reads with human data removed have been deposited in the European Nucleotide Archive (ENA) under ENA project accession number PRJEB62473.

### Authors’ contributions

Conceptualisation: E.C.-O. and A.C.D. Data curation: E.C.-O., B.P-S., M.W., C.A.N., K.M.M., S.H. and R.G. Formal analysis: E.C.-O. and Y.L. Funding acquisition: S.J.O’B. and N.A.C. Investigation: E.C.-O. and A.C.D. Project administration: E.C.-O., M.I.G., C.H.-F., S.J.O’B., N.A.C. and A.C.D. Resources: Y.L., B.P-S., J.C.D.H., C.A.N., C.H.-F. and A.C.D. Supervision: N.A.C. and A.C.D. Validation: E.C.-O. Visualisation: E.C.-O. and Y.L. Writing – original draft: E.C.-O., B.P-S., J.C.D.H., N.A.C. and A.C.D. Writing – review and editing: all authors.

## Supporting information

Supplementary File 1: Biom file containing k-mer counts and associated taxonomy from all INTEGRATE sequencing data.

Supplementary File 2: Coefficient value matrix generated using values extracted from Maaslin2 analysis to generate Figure 2 and Supplementary Figure 1

Supplementary File 3: Adjusted p-values generated using values extracted from Maaslin2 analysis to generate Figure 2 and Supplementary Figure 1.

Supplementary File 4: Genome coverage of diarrheal-associated Adenovirus in metagenomic and metatranscriptomic data.

Supplementary File 5: Traditional methods for pathogen identification by reference laboratories in the INTEGRATE study.

## Data Availability

Outputs from our analyses are available within the supplementary material of this manuscript. Illumina sequence reads with human data removed have been deposited in the European Nucleotide Archive (ENA) under ENA project accession number PRJEB62473.

https://www.ebi.ac.uk/ena/browser/view/PRJEB62473

## Acknowledgements

The authors thank the members of the INTEGRATE Consortium. The INTEGRATE Consortium investigators in the United Kingdom are Sarah J O’Brien (principal investigator), Frederick J Bolton, Rob M Christley, Helen E Clough, Nigel A Cunliffe, Susan Dawson, Elizabeth Deja, Ann E Durie, Sam Haldenby, Neil Hall, Christiane Hertz-Fowler, Debbie Howarth, Lirije Hyseni, Miren Iturriza-Gómara, Kathryn Jackson, Lucy Jones, Trevor Jones, K Marie McIntyre, Charlotte A Nelson, Lois Orton, Jane A Pulman, Alan D Radford, Danielle Reaves, Helen K Ruddock, Darlene A Snape, Debbi Stanistreet, Tamara Thiele, Maya Wardeh, David Williams, and Craig Winstanley (University of Liverpool), Kate Dodd (NIHR Clinical Research Network: North West Coast), Peter J Diggle, Alison C Hale, Barry S Rowlingson (Lancaster University), Jim Anson, Caroline E Corless, Viki Owen (Royal Liverpool and Broadgreen University Hospitals NHS Trust), Malcolm Bennett (University of Nottingham), Lorraine Bolton, John Cheesbrough, Katherine Gray, David Orr, Lorna Wilson (Lancashire Teaching Hospitals NHS Foundation Trust), Andrew R Dodgson, Ashley McEwan (Manchester University NHS Foundation Trust), Paul Cleary, Alex J Elliot, Ken H Lamden, Lorraine Lighton, Catherine M McCann, Matthieu Pegorie, Nicola Schinaia, Anjila Shah, Gillian E Smith, Roberto Vivancos, Bernard Wood (PHE), Rikesh Bhatt, Dyfrig A Hughes (Bangor University), Rob Davies (APHA); Simon de Lusignan, Filipa Ferreira, Mariya Hriskova, Sam O’Sullivan, Stacy Shinneman and Ivelina Yonova (University of Surrey/Royal College of General Practitioners).

---

**Supplementary Figure 1:**
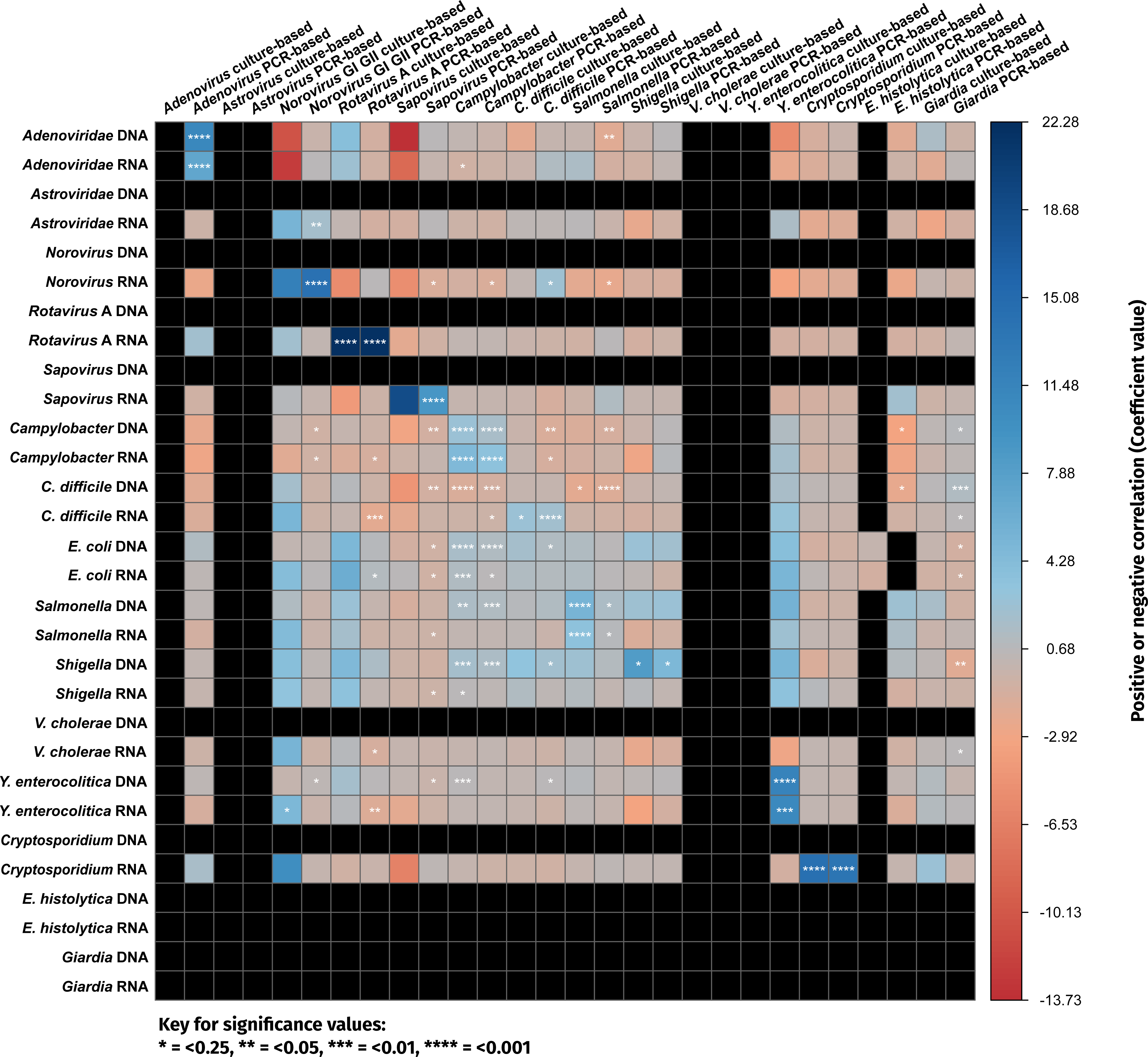
Complete overview of correlations observed between sequencing data and laboratory tests for major GI community pathogens in the United Kingdom (alternative to Figure 2).

