## Supplementary File 5: Traditional methods for pathogen identification by reference laboratories in the INTEGRATE study. for "Enhancing Infectious Intestinal Disease diagnosis through metagenomic and metatranscriptomic sequencing of over 1000 human diarrhoeal samples"

**BACTERIA**

#### Campylobacter

- Culture on *Campylobacter* selective media.
- Incubate at 42°C in a microaerophilic environment.
- **Appearance**: Grey-white colonies.
- **Gram Stain**: Gram-negative slender, curved or "seagull"-shaped bacilli.
- **Identification Tests**:
  - Oxidase positive (may be weak from charcoal media).
  - Confirm identification using MALDI-TOF, report species (e.g., *C. jejuni*).
  - Further characterisation using biochemical tests.

#### E. coli O157

- Culture on Sorbitol MacConkey (SMAC) agar.
- **Appearance**: Non-sorbitol fermenter (grey/colourless colonies).
- **Gram Stain**: Gram-negative bacilli.
- **Culture**:
  - Inoculate SMAC agar and incubate aerobically at 37°C for 48 hours.
- **Identification**:
  - Sub-culture suspect colonies on blood agar, incubate overnight.
  - Perform oxidase test; if negative, proceed to test using *E. coli* O157 latex test kit (Oxoid).
  - Test 10 NSF (non-sorbitol fermenter) colonies for O157.
  - Perform latex agglutination; ensure no reaction in control latex.
  - If agglutination in test latex, positive for *E. coli* O157.
  - Some strains may require emulsification or retesting with saline.
  - Confirm with API 20E or VITEK 2 (do not use MALDI-TOF unless by extraction).

#### Clostridioides difficile

- Routine culture not performed; test for toxins A and B using ELISA.
- If cultured, use Brazier’s medium.
- **Appearance**: Large colonies with yellow/green fluorescence under UV light.
- **Gram Stain**: Gram-positive sporing bacilli.

#### Salmonella

- Culture on XLD (Xylose Lysine Deoxycholate) and *Salmonella* selective media.
- **Appearance**: Pink colonies with or without black centres on XLD.
- **Gram Stain**: Gram-negative bacilli.
- **Identification**:
  - Identify suspicious colonies with MALDI-TOF.
- **Sensitivity Testing**:
  - Perform sensitivity tests on isosensitest agar.
  - Test against the "Gram-negative ring"; for *Salmonella*, also test against Naladixic Acid (30 μg).
  - For Naladixic Acid-resistant isolates, perform a Ciprofloxacin E-test.
- **Serology**:
  - Perform slide agglutination using *Salmonella* O, H, and Vi antisera.
  - If Vi antigen is positive, heat to remove and identify O group.
  - Further serology using Kauffmann White scheme to identify O and H antigens.

#### Shigella

- Culture on XLD and *Shigella* selective media.
- **Appearance**: Pink colonies without black centres on XLD.
- **Gram Stain**: Gram-negative bacilli.
- **Identification**:
  - Confirm suspicious colonies with MALDI-TOF.
- **Sensitivity Testing**:
  - Perform sensitivity tests on isosensitest agar.
  - Test against the "Gram-negative ring."
- **Serology**:
  - Identify serotypes using slide agglutination with specific O antisera.

#### Vibrio

- Culture on Colorex *Vibrio* Chromogenic Agar.
- **Appearance**:
  - *V. cholerae* and *V. vulnificus*: Pale blue colonies.
  - *V. parahaemolyticus*: Mauve colonies.
- **Gram Stain**: Gram-negative, comma-shaped bacilli.
- **Identification**:
  - Sub-culture on blood agar, incubate at 37°C overnight.
  - Oxidase positive.

#### Yersinia enterocolitica

- Culture on *Yersinia* Selective Agar (CIN agar) at 30°C for 48 hours.
- **Appearance**: Red colonies with transparent border ("bull’s eye" appearance).
- **Gram Stain**: Gram-negative bacilli.
- **Identification**:
  - Oxidase negative.
  - Confirm with MALDI-TOF.
- **Sensitivity Testing**:
  - Perform sensitivity testing on isosensitest agar.

**VIRUSES**

#### Method / Instructions for Performance of the Examination (Norovirus Focus)

#### Sample Preparation

1. **General Sample Collection for Viruses**

- Collect stool or vomit samples (minimum of 0.5ml).
- Store samples at +4°C until processing.
- Prepare samples using S.T.A.R buffer before nucleic acid extraction.

2. **Norovirus-Specific Sample Preparation**

- Wear gloves and work inside a Class 1 biosafety cabinet.
- Make a 20% solution of feces or vomit in S.T.A.R. Buffer (prepare at least 1ml).
- Vortex thoroughly and spin at 5000g for 5 minutes.
- Remove the supernatant into a fresh unskirted 2ml tube labeled with a barcode.
- Pipette 400µl into a 2ml unskirted tube if using the QIAcube for extraction.
- Use the unskirted barcoded tube if using the QIAsymphony for extraction.
- Store the remainder of the supernatant at -70°C after processing.

#### Nucleic Acid Extraction

1. **General Virus Extraction**

- Prepare a worklist of samples/controls to be extracted.

2. **QIAcube Virus Extraction**

- Use the MinElute Virus kit for nucleic acid extraction and elute into 60µl using the Large Body Fluid Standard Lysis Protocol.
- Add 1µl of Liv 10⁻⁴ MS2 phage per sample to the lysis buffer.

3. **QIAsymphony Virus Extraction**

- Use the Complex 400 V4 Default IC Protocol (see QIAgen QIAsymphony SP/AS DMS User Manual).
- Ensure that the carrier RNA solution containing MS2 phage is loaded.

4. **Sample Transfer for Norovirus**

- Transfer the 400µl supernatant into a barcoded unskirted 2ml tube for the QIAsymphony or QIAcube.

#### Master Mix Preparation

- In a clean room, prepare master mix using the ABI Taqman Virus One-Step RT-PCR kits.
- After centrifugation, pipette the master mix up and down several times before dispensing into the LC480 plate.
- Transfer the master mix tubes to the cool box.
- Store reagents at -20°C, avoiding multiple freeze-thaw cycles.
- In the PCR preparation enclosure, aliquot 5µl of master mix into the bottom of the appropriate wells in an LC480 96-well reaction plate (kept in a cool box).
- Add 5µl of sample or control to each corresponding well as per the worklist.
- Seal the plate and transfer it to the amplification area.

#### LightCycler Setup

- Prepare a LightCycler run worklist with sample numbers and technician information.
- Transfer prepared master mix to LC480 multiwell plates.
- Select "New experiment from Template" and choose the Gastro Taqman Fast protocol.
- Centrifuge the 96-well plate to collect fluids at the bottom (either 150xg for 1 minute or 1000xg for 1 second).
- Load the plate into the LC480 and start the run.
- Sample details may be entered once the run begins.

#### PCR Program

**LC480 Gastro Taqman Fast Run Protocol**

| Protocol | Temperature (°C) | Time (seconds) | Fluorescence Optics |
| --- | --- | --- | --- |
| RT | 50 | 300 |  |
| Denat | 95 | 20 |  |
| 45 Cycles | 95 | 3 |  |
|  | 58 | 45 | On |
| Cool | 40 | 30 |  |

#### Data Analysis

- Upon completion of the PCR run, examine amplification plots for each sample.
- Analyse fluorescence channels as follows:
  - **FAM** for Rotavirus.
  - **LC610** for Adenovirus.
  - **LC640** for Astrovirus.
  - **HEX** for Sapovirus.

Confirm results using internal quality controls, ensuring valid MS2 control signals.

2. **Norovirus-Specific Data Analysis**

- Apply the correct colour compensation: **NoV 3plx FAM-G2 HEX-G1 Cyan 500-MS2**.
- Manually inspect amplification plots for each sample and print the run report.
- Analyse the following channels:
  - **FAM** for Norovirus Genotype 2.
  - **HEX** for Norovirus Genotype 1.
  - **Cyan 500** for the MS2 internal control (IC).
- A positive result for **Norovirus Genotype 1** must have a CT <30 or HEX fluorescence >1.
- Ensure MS2 internal control signals are detected in the Cyan 500 channel for all negative samples (to rule out PCR inhibition).
- Consult with the Clinical Virologist for confirmation of results.

**PROTISTS**

#### Cryptosporidium

- Prepare and air-dry a smear of faeces.
- Fix smear by flame.
- Stain with Auramine Phenol stain for 10 minutes.
- Examine under a fluorescence microscope for yellow-green fluorescence.
- Confirm positive results with a control smear.

#### Giardia

- Use the Evergreen Faecal Parasite Concentrator system.
- Add 9 ml of 10% Formalin to a flat-bottomed tube.
- Add stool sample and mix.
- After 30 minutes, add 3 ml of Ethyl acetate and shake.
- Centrifuge at 500 x g for 10 minutes.
- Examine sediment for ova, cysts, and parasites.
- Use Lugol's iodine for small cysts like *Endolimax nana* and *Chilomastix mesnili*.
- Report findings.
